# Validity of a novel excessive daytime sleepiness’ screening stool : the Yaoundé Sleepiness Score

**DOI:** 10.1101/2024.02.16.24302934

**Authors:** Massongo Massongo, Duthoit Louise, Balkissou Adamou Dodo, Ngah Komo Marie Elisabeth, Kuaban Alain, Fry Stéphanie, Pefura Yone Eric Walter, Mallart Anne

**Author notes:** Corresponding author : Massongo Massongo, Faculty of Medicine and Biomedical Sciences, University of Yaoundé 1, PO Box 1364, Yaoundé – Cameroon. These authors contributed equally to this work (Balkissou Adamou Dodo, Ngah Komo Marie Elisabeth, Kuaban Alain, Fry Stéphanie, Pefura Yone Eric Walter).

## Abstract

**Study objective:** This study aimed to assess the validity of the Yaoundé Sleepiness Scale (YSS), a new tool for excessive daytime sleepiness (EDS) screening, using Epworth Sleepiness Scale (ESS) as reference.

**Methodology:** A 6-item questionnaire was assessed for content validity (based on sleep physiology, face validity (by 4 sleep specialists), criterion and construct validity (in a cross-sectional study led in Lille University Hospital). For criterion validity, we used Pearson correlation, area under the receiver operator characteristics (AUROC) curve and a graphical method to find the YSS thresholds. We performed a simple linear regression to seek the association between YSS and EDS predictors for construct validity.

**Results:** A total of 566 patients (mean age = 53.1 ± 14.6 years, female = 47%) were enrolled. The mean YSS and ESS were 9.8 ± 4.7 and 9.1 ± 5.3, respectively. The Pearson correlation between YSS and ESS was 0.74% (p<0.0001). The AUROC curves (95% confidence interval, 95CI) for ESS-based EDS and severe EDS prediction were 0.856 (0.829 – 0.889) and 0.871 (0.829 – 0.913), respectively. The YSS thresholds for EDS and severe EDS were 9 and 15, respectively. The sensitivity and specificity of YSS were 92.3 (88.7 - 95.9)% and 60.6 (55.3 - 65.9)%. The YSS was positively associated with psychiatric conditions and psychotropic drugs use, and negatively with age.

**Conclusion:** We found a good criterion validity of the YSS compared with ESS. This questionnaire could be proposed as an alternative to ESS.

## INTRODUCTION

Excessive daytime sleepiness (EDS) is a major symptom in sleep medicine. It may lead to road and work accidents, cognitive decline, impaired professional and school output, social friction, as well as increased health care consumption in sleep apnea syndrome (1). EDS has been associated with a higher risk of high blood pressure (HBP), insulin resistance, and cardiovascular mortality (2,3). An accurate evaluation of this symptom is thus fundamental for baseline assessment and follow-up of sleep disorders, including accidental risk.

Objective tools such as multiple sleep latency tests (MSLT) and maintenance of wakefulness test (MWT) were developed in early 80s to measure EDS (4–7), but these are laboratory-based, not affordable for most of the patients and physicians. Therefore, easy-to-use tools have been proposed, the most known and used in clinical practice being the Epworth Sleepiness Scale (ESS), which evaluates the sleepiness risk in 8 different daily situations, relating to the usual way of life in recent times (8). However, some items of ESS are not designed to be universally answered, especially those that require some equipment (electricity supply and television set for item 2 = watching television), some habits or possibilities (owing or getting access to a car or public transport for item 4 = as a passenger in a car, and 5 = in a car while stopped for a few minutes in the traffic), and some abilities (being literate for item 1 = sitting and reading). This has been illustrated in some studies where 59 to 64% of the study population were unable to complete properly ESS, not only in elderly people without dementia (9,10), but also in young and urban adults (11).

We assessed the validity of a new EDS questionnaire, in which items are based on internal and physiological features rather than external ones, with the purpose to make it universally easy to use. Various aspects of validity were studied: face, content, criterion, and constructs. Normative values were also sought, using the ESS as reference. The ESS is coted 0 to 24, scores ˃ 10 and 16 indicate an EDS and a severe EDS respectively.

## METHODOLOGY

### Presentation of the Yaounde sleepiness scale (YSS)

The YSS is designed to be self-administered for healthy or ill adults aged 18 years and above, after a usual sleep night, excluding any obvious cause of insufficient sleep time. It is made up of 6 questions divided into 3 domains (see appendix)

- **Feeling of EDS** : 1 question scoring 0 to 4 points;
- **Circumstances in which EDS occurs** : 3 questions scoring 0 to 3 points each giving a total of 12 points;
- **Consequences of EDS** in terms of daily life disturbance (1 question, 0 – 4 points) and sleep latency (1 question, scoring 0 to 3)

The total score varies from 0 to 20, 0 representing the absence of EDS and 20 its highest level (Table 1).

**Table 1.**
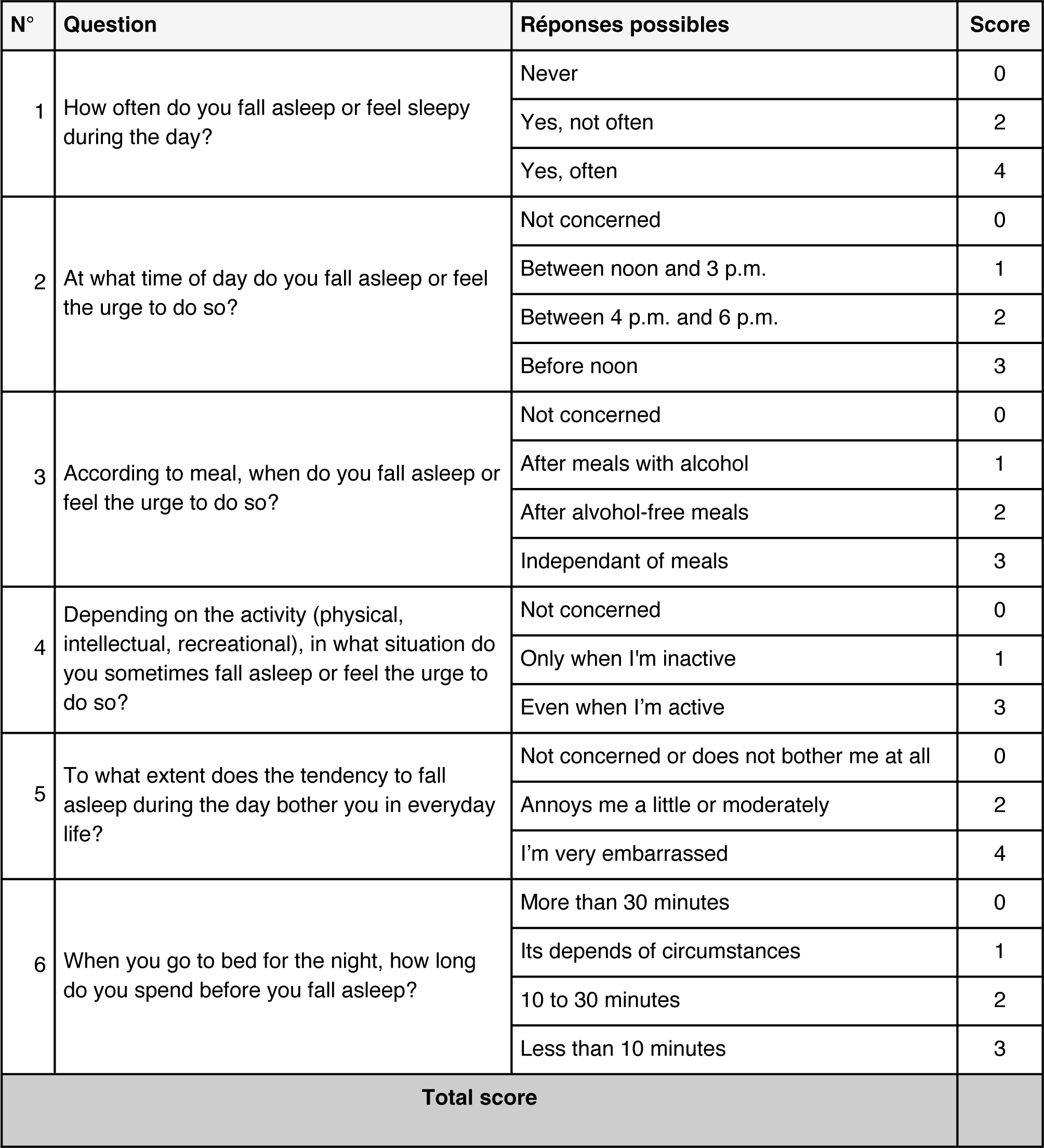
Presentation of the Yaoundé sleepiness scale, translated from the original version in French.

| N° | Question | Réponses possibles | Score |
| --- | --- | --- | --- |
| 1 | How often do you fall asleep or feel sleepy during the day? | Never | 0 |
|  |  | Yes, not often | 2 |
|  |  | Yes, often | 4 |
| 2 | At what time of day do you fall asleep or feel the urge to do so? | Not concerned | 0 |
|  |  | Between noon and 3 p.m. | 1 |
|  |  | Between 4 p.m. and 6 p.m. | 2 |
|  |  | Before noon | 3 |
| 3 | According to meal, when do you fall asleep or feel the urge to do so? | Not concerned | 0 |
|  |  | After meals with alcohol | 1 |
|  |  | After alcohol-free meals | 2 |
|  |  | Independent of meals | 3 |
| 4 | Depending on the activity (physical, intellectual, recreational), in what situation do you sometimes fall asleep or feel the urge to do so? | Not concerned | 0 |
|  |  | Only when I'm inactive | 1 |
|  |  | Even when I'm active | 3 |
| 5 | To what extent does the tendency to fall asleep during the day bother you in everyday life? | Not concerned or does not bother me at all | 0 |
|  |  | Annoys me a little or moderately | 2 |
|  |  | I'm very embarrassed | 4 |
| 6 | When you go to bed for the night, how long do you spend before you fall asleep? | More than 30 minutes | 0 |
|  |  | Its depends of circumstances | 1 |
|  |  | 10 to 30 minutes | 2 |
|  |  | Less than 10 minutes | 3 |
| Total score |  |  |  |

### Content validity

We used the available knowledge on sleep and awakening physiology, as well as clinical semiology, to assume the relevance of each item used in the YSQ, as well as the eligible population.

- **Population = 18 years old and above** : The adult pattern of sleep started from 18 years of age. The sleep physiology varies constantly from birth to 18 (12–15).
- **Question 1 = presence/frequency of feeling EDS**: Prior to its assessment, we first sought the presence of EDS felt by the subject. Most of the subsequent questions would depend on this feeling of EDS.
- **Question 2** = **time of occurrence in the day**. Due to the circadian clock, the daytime is not equally distributed regarding wakefulness. Wakefulnessis minimal at the end of the night (2 – 5 AM), decreased at the beginning of the afternoon (2 - 4 PM), and maximal at the end of the morning and evening (16). Thus, EDS will be even more important as it occurs during increased wakefulness periods, and vice versa.
- **Question 3 = relation to food intake**. The postprandial period is characterized by decreased wakefulness, due to a shift of the bloodstream towards the digestive system. Thus, feeling asleep far from a meal would be more significant than close to it.
- **Question 4 = relation to activity**. Physical, intellectual, or fun activities keep the nervous system awake. Feeling asleep during activity is therefore a sign of very significant EDS.
- **Question 5 = impact on daily life (professional, scholar, or social).** The more EDS will be harmful for the person, the more it should be considered severe
- **Question 6 = sleep latency**. Sleep latency is used to objectively measure EDS (MSLT). Its estimation can be a relevant item in a subjective score. The more it is short, the more EDS will be important.

### Face validity

The YSS was submitted to 2 chest physicians in Yaoundé with sleep disorders training and experience, as well as 2 experts from sleep laboratory of Lille University Hospital (LUH) for evaluation and approval. All of them subjectively approved the original draft.

### Criterion and external construct validity

To achieve these main issues, we conducted a study which aimed :

1. To seek a correlation between YSS with ESS (criterion validity)
2. To set a positivity threshold of the YSS in detecting daytime sleepiness (using ESS as gold standard) and assess its performance
3. To seek an association between YSS and risk factors for EDS (construct validity).

### Study design, setting, and population

This was a cross-sectional study which occurred in the respiratory department of Lille University Hospital (LUH), which is the reference hospital of the former Nord Pas-de-Calais Region, France.

From November 1, 2016 to June 30, 2017, literate adults admitted for a full night respiratory exploration were consecutively invited to participate, excluding those who were already on treatment with respiratory devices such as continuous positive airway pressure (CPAP) or noninvasive ventilation (NIV).

### Procedure

Primary data were collected during the daytime activities of the sleep explorations unit. Administrative data were already present in the electronic patient’s file. Patients were asked to fill a YSS as part of the usual files submitted at their entrance in the unit, including ESS, identification sheets and others. Anthropometric data were collected by the paramedical staff and the clinical observation was performed by the medical staff of the unit. The investigator of the present study, who was in his final year of training in pulmonology, collected data retrospectively from patients’ files, from 1^st^ to 30^th^ June 2017.

Baseline data included: age, gender, occupation, smoking status, weight category based on body mass index (BMI), categorized in normal/thin, overweight, and obesity.

The following conditions likely to cause EDS were sought: psychotropic drug consumption, alcohol, known neurologic or psychiatric disease, metabolic or endocrine disease, cardiovascular disease, non respiratory sleep disorder, history of skull trauma, sleep debt.

Completeness of the filling of the YSS and ESS was categorized as: empty (no items informed on), incomplete (at least one item not filled), or complete (all items filled).

The sleep respiratory exploration was either a respiratory polygraphy (RP) using Noxturnal software (ResMed Corp, San Diego, USA) and recording 4 main items (air flow, pulse rate, transcutaneous oxygen saturation of hemoglobin and thoracic abdominal movements); or full polysomnography (PSG) using the Medatec software (Medical Data Technology SPRL-BVBA, Braine-Le-Chateau, Belgium) that recorded at least three supplementary measurements (eye electromyogram, electroencephalogram and leg electromyogram). The choice between those two methods was done in advance by the staff, depending on the underlying or suspected condition.

### Data analysis

We used Open Epi software version 3 to estimate the sample size. Assuming that 85 +/- 5% of patients having ESS-based EDS would have YSS-based EDS, we needed 196 patients to properly estimate this proportion with à 95% confidence interval. Knowing that about half of patients attending the unit are often sleepy with ESS, and anticipating a 10% non-respondent rate, the final needed sample size was 431. We processed and analyzed our data using R, version 4.0.5 for Windows (SAS Institute, Cary, NC, USA).

### Baseline data

Categorical data were expressed as counts (%), and numerical ones as mean ± standard deviation when normally distributed or median (1^st^, 3^rd^ quartiles) otherwise.

### Criterion validity

We used the graphic method and/or the Shapiro test to check the normal distribution of numerical variables. We used the Pearson correlation coefficient to assess the correlation between YSS and ESS as numerical variables, in the whole sample and in subgroups. We assumed a significant coefficient ˃ 0.7. We estimated the area under the curve (AUC) and 95% confidence interval (95CI) of the receiver operator characteristics (ROC) curve to assess YSS ability to predict ESS-based EDS. We expected the lower limit of 95CI ˃ 0.8.

For the diagnosis threshold and performance of YSS in predicting ESS-based EDS, we did not find a consensus or validated method in the literature. We then used a graphical method, based on the proportion of participants presenting an ESS-based ESD, among the 21 values of YSS (0 to 20), ranged in an increasing order. We assumed that each sudden increase in that proportion from one value to the following one, would indicate a new category of subjects regarding the risk of EDS. The sensitivity and specificity of the resulting threshold were also calculated.

### Construct validity

The association between YSS and factors likely to influence EDS was sought using simple linear regression. The explanatory variables were age, the presence of sleep disordered breathing (defined as an apnea hypopnea index on polysomnography ≥ 5/hour), weight, and the conditions likely to cause EDS, listed above (procedure section).

For all these analysis, the threshold for rejecting the null hypothesis (p-value) was 0.05.

### Ethics

The present study was approved by the Ethical Board of Lille University Hospital. Each participant received an information sheet and gave an oral consent prior to his enrollment. Since this was an observational study, the ethics board did not make written consent mandatory. Study-related data were collected anonymously.

## RESULTS

### Baseline data

A total of 566 patients were included in the study. Their mean (SD) age was 53.1 (14.6) years, and half of them were male. About 2/3 had less than one-hour physical activity per week and more than 4/5 were overweight or obese. Among the participants, 533 (94.2%) and 562 (97.3%) filled the YSS and the ESS, respectively. The completeness was comparable between the 2 questionnaires. SDB was found in 80% of patients, and moderate to severe sleep disordered breathing (MS-SDB) in more than half of them. These data are detailed in Table 2.

**Table 2.** baseline data of the patients enrolled in YSS validation study, Lille, 2017 - 2018. N = 566.

| Sections | Variables | Sample size | Values* |
| --- | --- | --- | --- |
| Sociodemographic data | **Age (years), numerical** | 566 | 53.1 (14.6) |
|  | **Age (years), categorical** | 566 |  |
|  | 18 - 39 |  | 118 (20.8) |
|  | 40 - 64 |  | 312 (55.1) |
|  | 65 - 79 |  | 121 (21.4) |
|  | 80+ |  | 15 (2.7) |
|  | **Sex** | 566 |  |
|  | Female |  | 268 (47.0) |
|  | Male |  | 298 (53.0) |
|  | **Occupation** | 461 |  |
|  | Active |  | 236 (51.2) |
|  | Inactive |  | 225 (48.8) |
|  | **Physical activity** | 401 |  |
|  | > 1hour/Week |  | 131 (32.7) |
|  | ≤ 1hour/Week |  | 270 (67.3) |
|  | **Body mass index (Kg/m<sup>2</sup>)** | 553 | 30.4 (7.5) |
|  | **Weight category** | 553 |  |
|  | Normal or thin |  | 129 (23.3) |
|  | Overweight |  | 163 (29.5) |
|  | Obese |  | 261 (47.2) |
| **Yaounde sleepiness scale (YSS)** | Completeness | 533 | 514 (96.4) |
|  | Mean score |  | 9.8 (4.7) |
| **Epworth sleepiness scale (ESS)** | Completeness | 562 | 547 (97.3) |
|  | Mean score |  | 9.1 (5.3) |
| | Sleepiness (ESS ≥ 11) | | 217 (38.3) |
| | MS sleepiness (ESS ≥ 16) | | 75 (13.2) |
| **EDS\*\* predisposing conditions factors** | Alcohol consumption | 268 | 18 (6.7) |
|  | Tobacco smoking | 266 | 59 (21.8) |
|  | Psychotropic drugs | 268 | 99 (36.7) |
|  | Neurologic | 268 | 59 (21.8) |
|  | Psychiatric | 268 | 59 (21.8) |
|  | Endocrine and metabolic | 268 | 123 (45.6) |
|  | Cardiovascular | 268 | 123 (45.6) |
|  | Sleep debt | 267 | 28 (10.4) |
| **Polysomnography results** |  | 270 |  |
|  | Apnea hypopnea index |  | 22.8 (20.2) |
|  | Sleep disordered breathing |  | 216 (80.0) |
|  | MS_SDB*** |  | 151 (55.9) |

\* *Categorical data are expressed as counts (%) and continuous data as means (SD)*
\*\* *EDS : excessive daytime sleepiness*
\*\*\**MS\_SDB: moderate to severe sleep disordered breathing*

### Criterion validity

#### Yaounde and Epworth scale correlation

We found a significant correlation between YSS and ESS, with a Pearson coefficient (95% confident interval, P value) of 0.74 (0.70 – 0.77, p<0.0001). The resulting scatter plot is presented in Figure 1.

**Figure 1.**
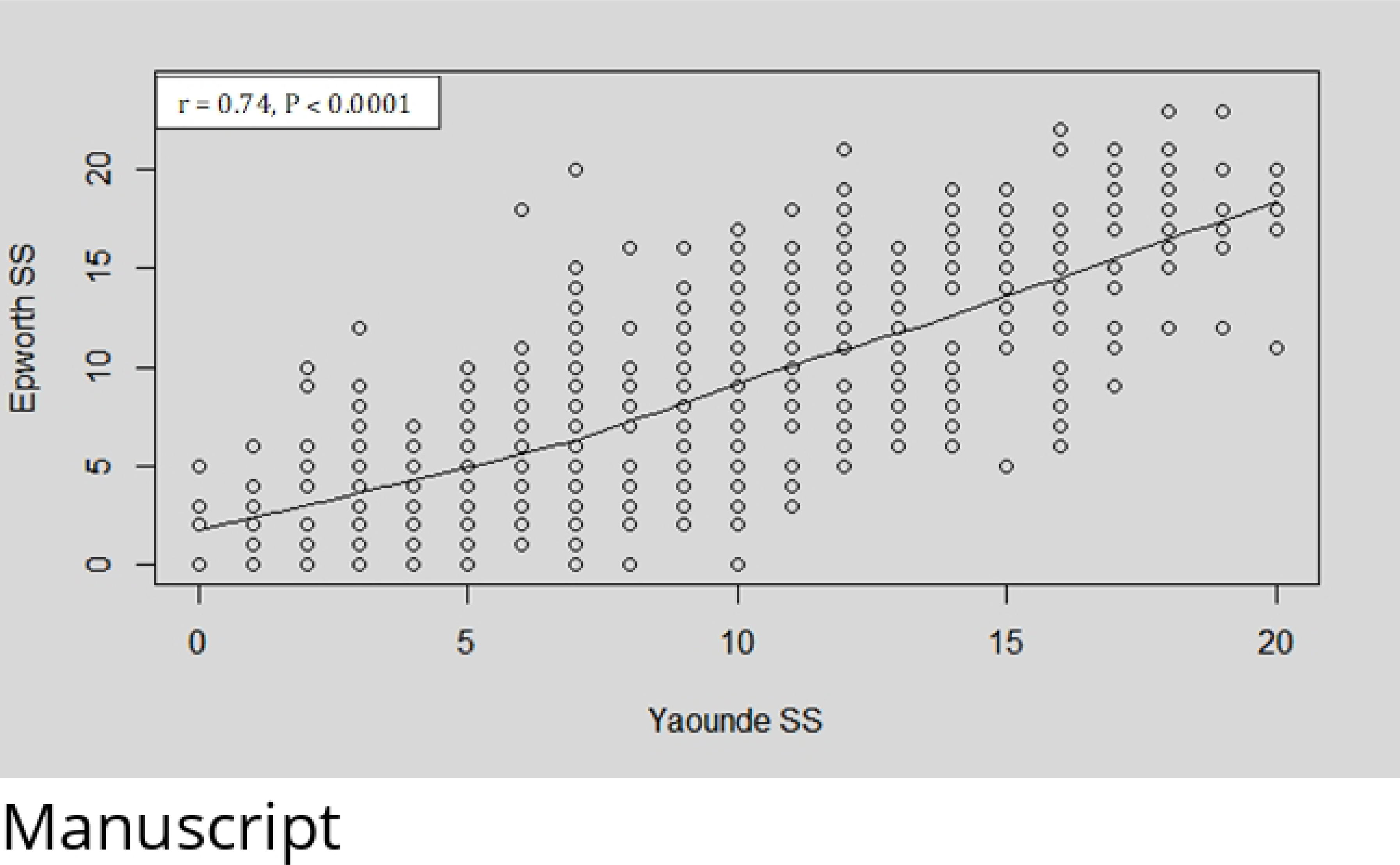
**Scatter plot presenting the correlation between Yaoundé sleepiness score and Epworth sleepiness score.**

The YSS mean ± SD was significantly higher in ESS-based EDS+ than EDS- (13.3 ± 3.6 vs 7.4 ± 3.9, p<10^-15^). A similar result was found for severe EDS (15.2 ± 3.5 vs 8.8 ± 4.2, p<10^-15^).

This correlation was confirmed in different subgroups with respect to age, occupation, or physical activity, although the Spearman coefficient was lesser in the > 80 years group; as shown in Table 3.

**Table 3.** Pearson correlation coefficient between Yaoundé and Epworth sleepiness scores with respect to age, occupation, and physical activity.

| Population group |  | Sample size | Pearson coefficient | P-value |
| --- | --- | --- | --- | --- |
| **Global sample** |  | 534 | 0.74 | <0.0001 |
| **Age groups (years)** | 18 - 39 | 118 | 0.78 | <0.0001 |
|  | 40 - 64 | 312 | 0.70 | <0.0001 |
|  | 65 – 79 | 121 | 0.70 | <0.0001 |
|  | ≥ 80 | 15 | 0.63 | 0.0300 |
| **Occupation** | Active | 236 | 0.75 | <0.0001 |
|  | Inactive | 261 | 0.72 | <0.0001 |
| **Physical activity** | Active | 270 | 0.75 | <0.0001 |
|  | Inactive | 131 | 0.73 | <0.0001 |

#### YSS and Epworth-based sleepiness prediction

The AUROC (95CI) for ESS-based EDS prediction by YSS was 0.856 (0.829, 0.889). The related ROC curve is presented in Figure 2.

**Figure 2.**
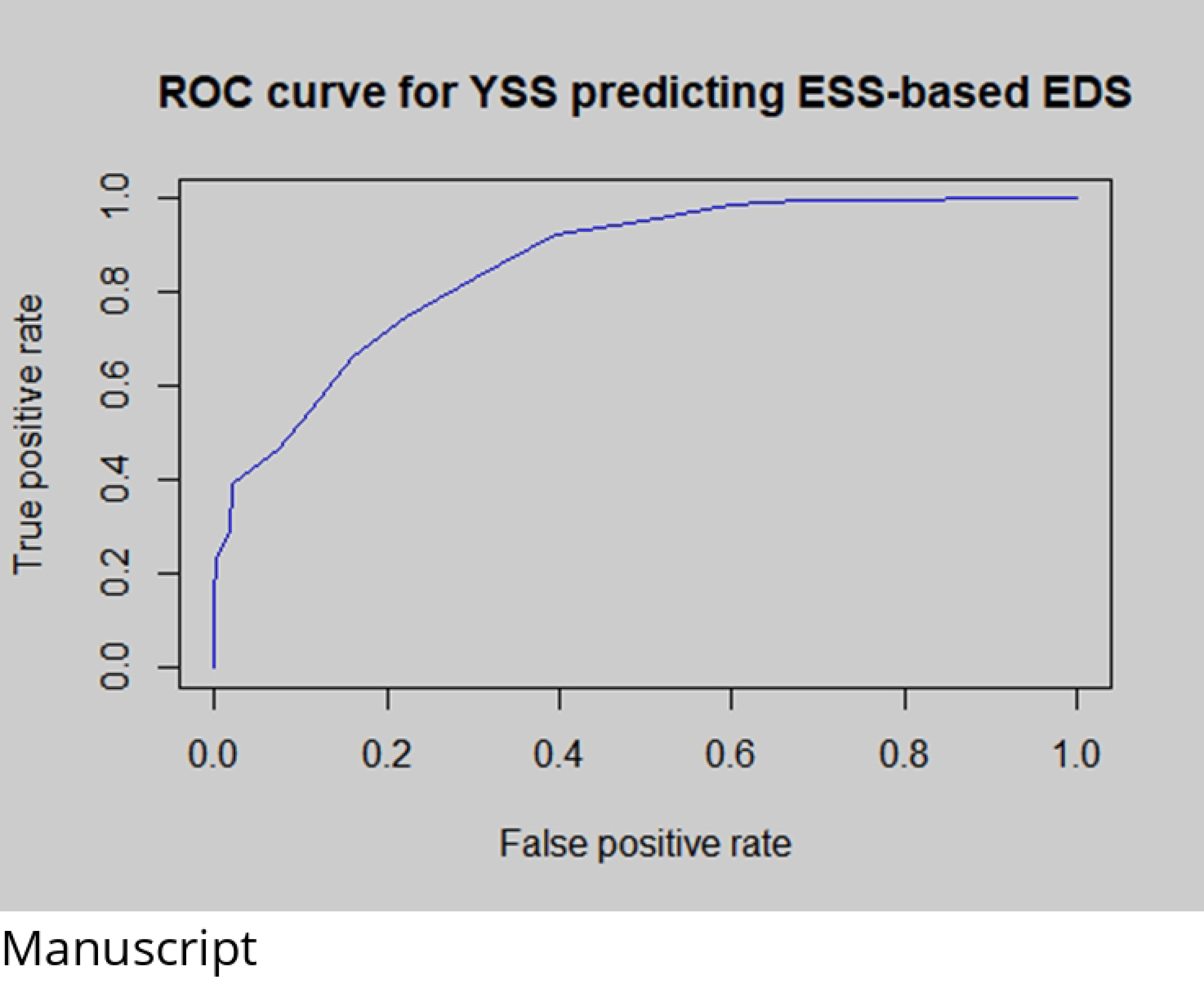
**ROC curve for Epworth based sleepiness prediction by Yaounde sleepiness scale.**

The AUROC (95CI) for severe ESS-based EDS prediction by YSS was 0.871 (0.829, 0.913). The related ROC curve is presented in Figure 3.

**Figure 3.**
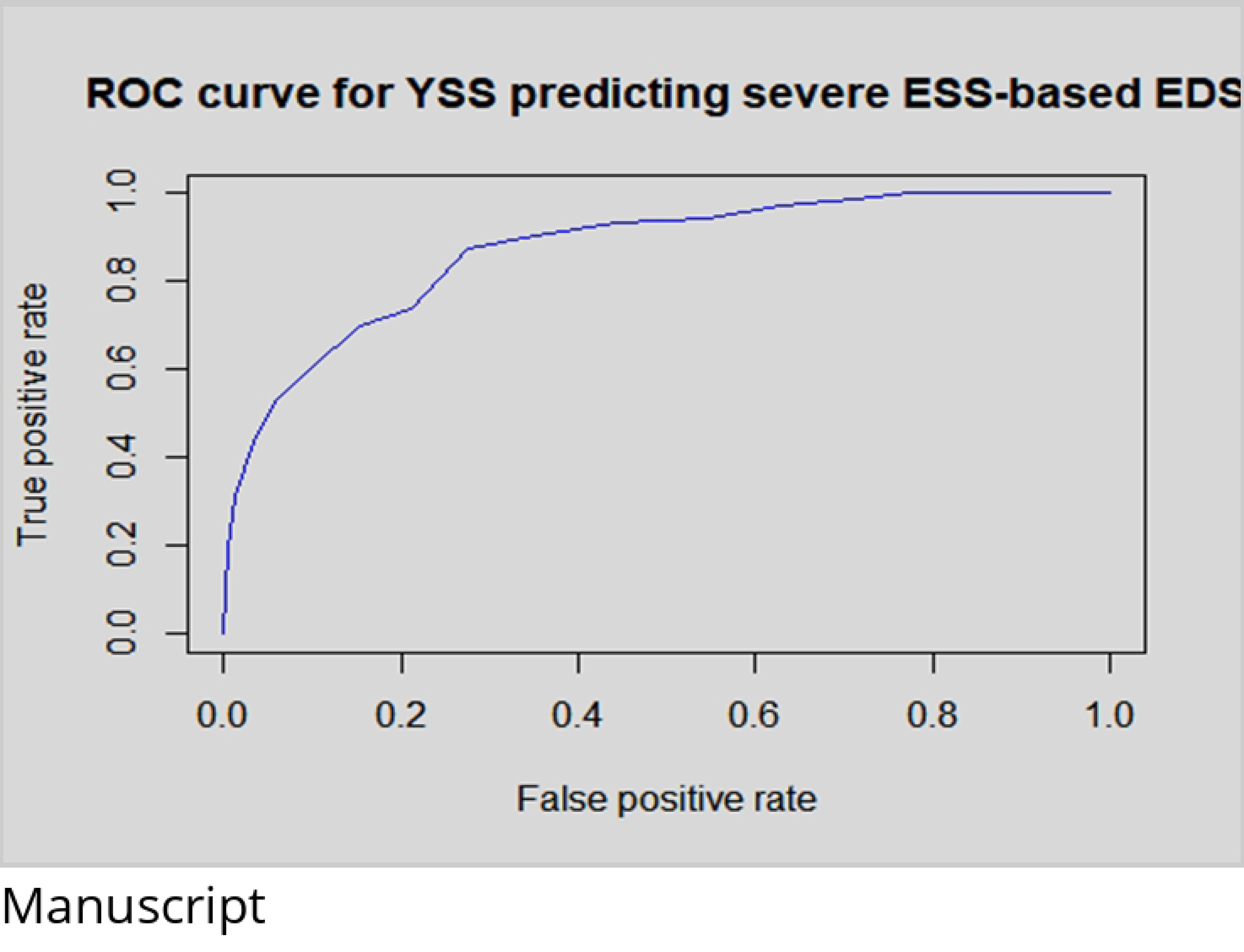
**ROC curve for Epworth based severe sleepiness prediction by Yaounde sleepiness scale.**

#### YSS thresholds and diagnostic performance

Using the graphical method described above, we obtained two sudden increases in the proportion of participants with significant ESS. These occurred at thresholds 9 and 15 (Figure 4), which were then considered as defining YSS-based EDS and severe EDS, respectively.

**Figure 4.**
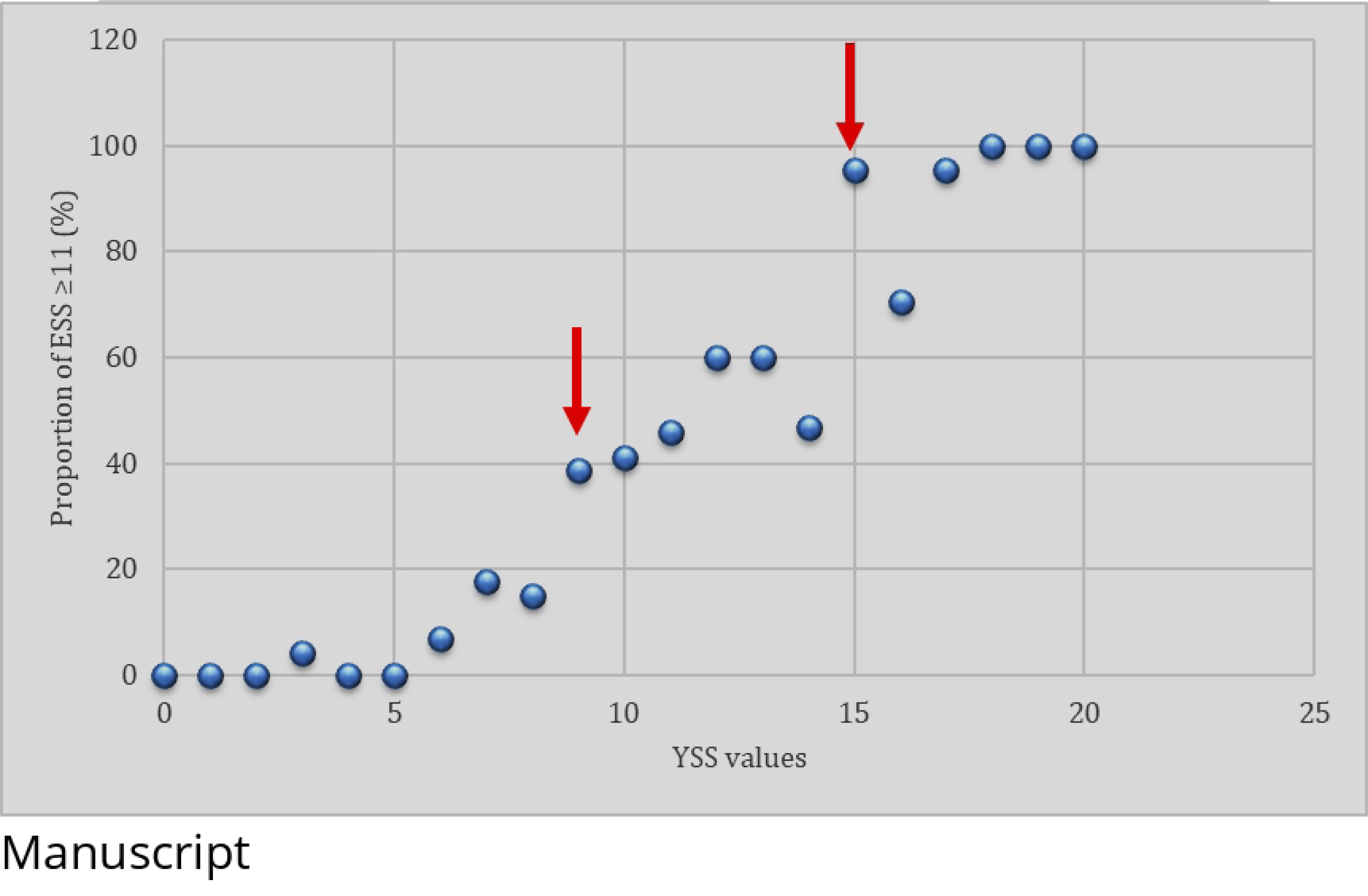
**Proportion of participants with significant Epworth sleepiness score among each group of Yaoundé sleepiness scores. Arrows indicate the sudden increase in proportion.**

Using these definitions to assess the diagnosis performance of YSS with ESS as the gold standard, we obtained the data presented in Tables 4 and 5. As result, the YSS sensitivity and specificity (95CI) were, respectively, 92.3 (88.7 - 95.9)% and 60.6 (55.3 - 65.9)% for EDS diagnosis; and 60.3 (49.0 - 71.5)% and 90.2 (87.5 - 92.9)% for severe EDS diagnosis.

**Table 4.**
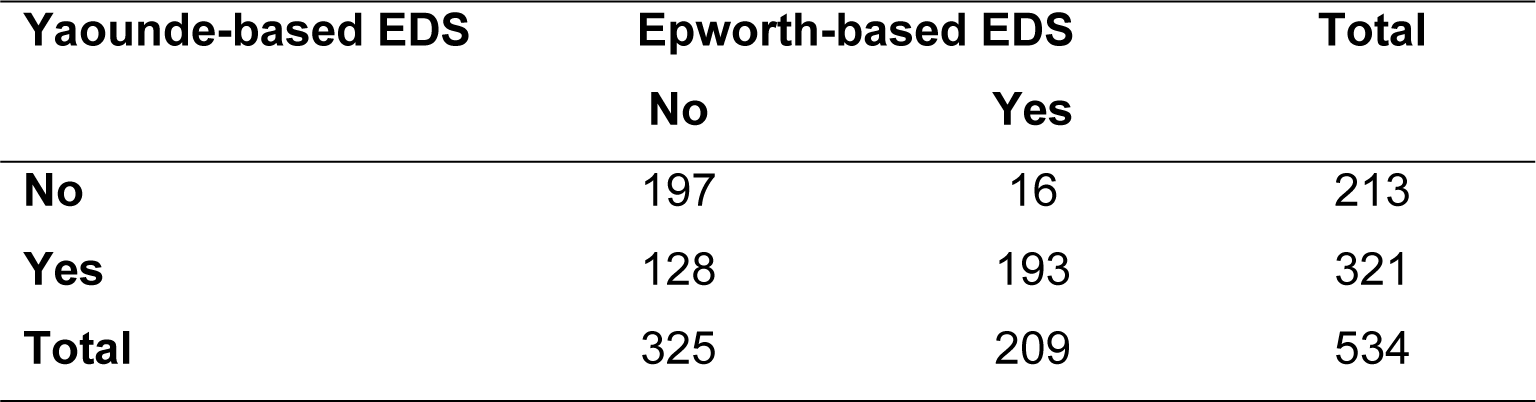
Contingency table of YSS-threshold 9 and ESS-threshold 11 for EDS diagnosis.

| Yaounde-based EDS | Epworth-based EDS |  | Total |
| --- | --- | --- | --- |
|  | No | Yes |  |
| No | 197 | 16 | 213 |
| Yes | 128 | 193 | 321 |
| Total | 325 | 209 | 534 |

**Table 5.** Contingency table of YSS-threshold 15 and ESS-threshold 16 for severe EDS diagnosis.

| Yaounde-based severe EDS | Epworth-based severe EDS |  | Total |
| --- | --- | --- | --- |
|  | No | Yes |  |
| No | 416 | 29 | 445 |
| Yes | 45 | 44 | 89 |
| Total | 461 | 73 | 534 |

### Construct validity

Using linear regression, we found that the Yaounde sleepiness score significantly increased with the presence of psychiatric conditions, the use of psychotropic drugs, and the presence of EDS ground as defined above; while it decreased with age. The was a non-significant increase with neurologic conditions and sleep depth. Alcohol consumption and weight category did not influence the score (Table 6).

**Table 6.**
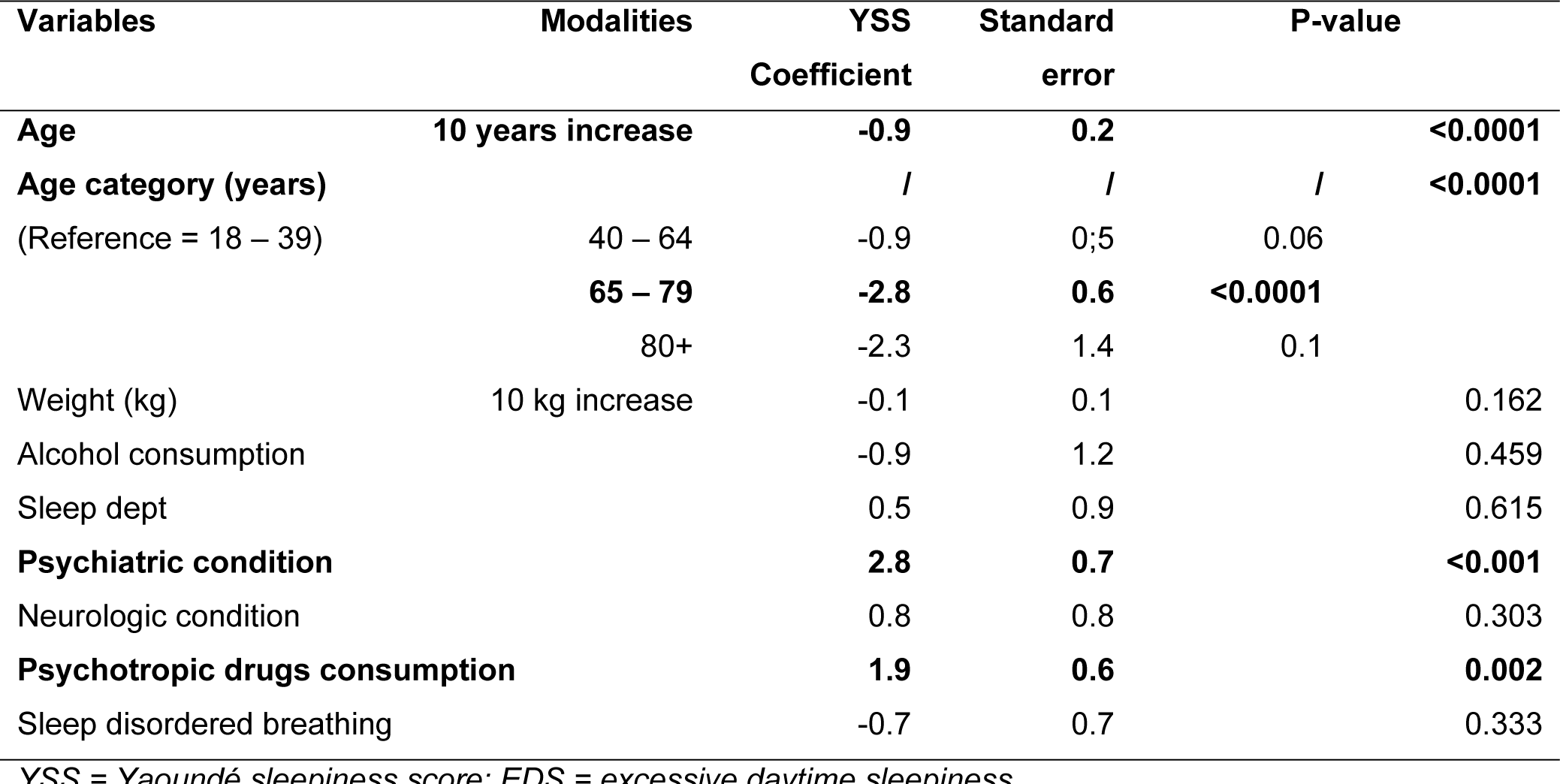
Results of linear regression modeling the relationship between YSS and some variables likely to influence EDS.

## DISCUSSION

The present study aimed to validate a new EDS assessment tool, using ESS as the gold standard. Items used in this questionnaire are related to sleep physiology and can be easily informed by any person, regardless his social, environmental, or economic status. The questionnaire has been approved by experts in the sleep field. For the criterion validity, a significant correlation has been found with the ESS, and the YSS showed a good area under the ROC (AUROC) curve for EDS prediction. Thresholds for EDS and severe EDS have been estimated. Using those thresholds, YSS appeared to be very sensitive and less specific for EDS detection globally, and conversely for severe EDS. Significant association has been found between YSS and some conditions likely to cause EDS, suggesting an acceptable construct validity.

We tried our best to follow the main steps in the validation process of a diagnostic tool. However, a non-expert population has not been involved in the face validity step. This would have provided an insight into the target population. We found comparable completeness rate between the two scales, suggesting the absence of supplementary difficulties for YSS, although it was totally new for patients as well as care givers. However, the detailed analysis of the responses, especially for ESS, revealed that some participants answered improperly to some items : for example, concerning sleepiness in a car as a driver, for a patient who did not have a car nor drove. This was a potential source of bias. The real life condition and consecutive recruitment contributed to a reduced risk of selection bias.

The positive correlation between YSS and ESS suggests a real capacity of YSS in detecting subjective EDS. The high values of the area under the ROC curve reinforce this feeling, both for global and severe daytime sleepiness prediction. The good sensitivity we obtained for global EDS detection supports the relevance of our lower YSS threshold and can reinforce the YSS as a screening tool, for which a low specificity is not detrimental. Interestingly, the specificity of YSS for severe EDS detection was much higher, making it useful and relevant as a diagnostic tool, since severe EDS is known to be associated with a worse outcome. In this perspective, we can consider a double advantage for YSS.

The absence of an association between YSS and SAS was not surprising, since this is frequent in the literature on ESS, as EDS is not specific to sleep apnea or sleep disordered breathing, but may be the consequence of various conditions. Rather, ESS has been found to be associated with AHI and oxygen desaturation index (ODI) among OSA patients (17,18), probably because of the related arousals. Reports on the association between EDS and other features likely to cause EDS, in the absence of a given disease, are variable. Concerning the use of alcohol as a predictor of EDS, Filomeno recently found a strong association between ESS and alcohol intake, in a population quite different from our own (Japanese male commercial truck drivers aged 43 years or more) (19), while this association was less important in another study among patients suspected of having obstructive sleep apneas (20). Psychiatric conditions and use of psychotropic drugs have been widely described as EDS determinants, especially depression (20–24). We could also expect a correlation between YSS and weight or body mass index, as shown by Bixler et al. (21) or Koutsourelakis et al. (20) a few years ago, but this did not occur in our study. Interestingly, we found a decrease of YSS with increasing age. This is consistent with knowledge on sleep physiology. It has been shown that the total sleep time needed decreases with age (25). Thus, under the same conditions, older people will be expected to feel less sleepiness than younger. This has been observed by Stenuit and Kerkhofs among ladies after sleep deprivation (26). The association of YSS with some of these features suggests a valuable argument for construct validity.

These results support the efficacy of YSS in predicting EDS and the relevance of it items. As such, it may be proposed in the areas were using the ESS may lead to systematic bias, due to its items which are not all applicable to everyone, as described in the Background section. Low-and-middle income contries (LMICs) are particularly concerned by this issue, and this should not be neglected, since non communicable diseases and sleep disorders are increasing in these countries. We also noticed that even in developed countries, some populations, especially the elderly, are concerned by difficulties in filling ESS (9,10), reinforcing the need for an easier and more universal tool for EDS screening.

However, the process is not yet completed before the generalization of YSS. Its reliability has not yet been demonstrated, this should include reproducibility and change sensitivity studies, which are ongoing. It also needs validity studies among target populations in LMIC. Ideally we should validate our subjective screening tool against an objective assessment of sleepiness. However the tools for the objective assessment of sleepiness were not readily available in our settings at the stud yset up. As a result, we decided to first compare our screening tool with the widely used and universally accepted Epworth Sleepiness scale to see how our tool faired in comparison to the latter. After obtaining data supporting the new screening tool, we now plan to test the screening tool against an objective assessment sleepiness test.

## CONCLUSIONS

This study demonstrated a strong correlation between the Yaoundé and Epworth sleepiness scores. The YSS was a good predictor of ESS-based daytime sleepiness. The YSS thresholds to define global (ESS 11) and severe (ESS 16) EDS were 9 and 15, respectively. The YSS increased with psychiatric conditions and use of psychotropic drugs, while it decreased with age. These results suggest a good criterion validity and acceptable construct validity. The YSS could be proposed as an alternative of ESS, but confirmatory data (especially based on objective sleepiness tests) and reliability studies are still needed.

## Data Availability

Data supporting the conclusions of this study can be accessed upon reasonable demand, by email to

## ACKNOWLEDGEMENTS

We sincerely thank the staff of Respiratory clinic of Lille University Hospital, as well as all participants.

## REFERENCES

1. Ronksley PE, Hemmelgarn BR, Heitman SJ, Flemons WW, Ghali WA, Manns B, et al. Excessive daytime sleepiness is associated with increased health care utilization among patients referred for assessment of OSA. Sleep. 2011;34(3):363–70.

2. Gooneratne NS, Richards KC, Joffe M, Lam RW, Pack F, Staley B, et al. Sleep disordered breathing with excessive daytime sleepiness is a risk factor for mortality in older adults. Sleep. 2011;34(4):435–42.

3. Xie J, Sert Kuniyoshi FH, Covassin N, Singh P, Gami AS, Chahal CAA, et al. Excessive Daytime Sleepiness Independently Predicts Increased Cardiovascular Risk After Myocardial Infarction. J Am Heart Assoc. 2018;19:7(2).

4. Carskadon MA, Dement WC. The multiple sleep latency test: what does it measure? Sleep. 1982;5 Suppl 2:S67–72.

5. Carskadon MA, Dement WC, Mitler MM, Roth T, Westbrook PR, Keenan S. Guidelines for the multiple sleep latency test (MSLT): a standard measure of sleepiness. Sleep. 1986;9(4):519– 24.

6. Mitler MM, Gujavarty KS, Browman CP. Maintenance of wakefulness test: A polysomnographic technique for evaluating treatment efficacy in patients with excessive somnolence. Electroencephalogr Clin Neurophysiol. 1982;53(6):658–61.

7. Doghramji K, Mitler MM, Sangal RB, Shapiro C, Taylor S, Walsleben J, et al. A normative study of the maintenance of wakefulness test (MWT). Electroencephalogr Clin Neurophysiol. 1997;103(5):554–62.

8. Johns MW. A new method for measuring daytime sleepiness: the Epworth sleepiness scale. Sleep. 1991;14(6):540–5.

9. Onen F, Moreau T, Gooneratne NS, Petit C, Falissard B, Onen SH. Limits of the Epworth Sleepiness Scale in older adults. Sleep Breath. 2013;17(1):343–50.

10. Frohnhofen H, Popp R, Willmann V, Heuer HC, Firat A. Feasibility of the Epworth Sleepiness Scale in a sample of geriatric in-hospital patients. J Physiol Pharmacol. 2009;60 Suppl 5:45–9.

11. Massongo M, Bitchong CE, Bassogbag R, Ze EA. The Epworth Sleepiness Scale’s Completeness Difficulties in an Urban Cameroonian Population. Health Sci Dis [Internet]. 2020 May 20 [cited 2021 Jun 3];21(6). Available from: http://hsd-fmsb.org/index.php/hsd/article/view/2052

12. Anders TF, Carskadon MA, Dement WC. Sleep and sleepiness in children and adolescents. Pediatr Clin North Am. 1980;27(1):29–43.

13. Carskadon MA, Harvey K, Duke P, Anders TF, Litt IF, Dement WC. Pubertal changes in daytime sleepiness. Sleep. 1980;2(4):453–60.

14. Scholle S, Beyer U, Bernhard M, Eichholz S, Erler T, Graness P, et al. Normative values of polysomnographic parameters in childhood and adolescence: quantitative sleep parameters. Sleep Med. 2011;12(6):542–9.

15. Galland BC, Taylor BJ, Elder DE, Herbison P. Normal sleep patterns in infants and children: a systematic review of observational studies. Sleep Med Rev. 2012;16(3):213–22.

16. Dijk DJ, Duffy JF, Riel E, Shanahan TL, Czeisler CA. Ageing and the circadian and homeostatic regulation of human sleep during forced desynchrony of rest, melatonin and temperature rhythms. J Physiol. 1999;516(2):611–27.

17. Rey de Castro J, Rosales-Mayor E. Clinical and polysomnographic differences between OSAH patients with/without excessive daytime sleepiness. Sleep Breath. 2013;17(3):1079–86.

18. Mediano O, Barceló A, de la Peña M, Gozal D, Agustí A, Barbé F. Daytime sleepiness and polysomnographic variables in sleep apnoea patients. Eur Respir J. 2007;30(1):110–3.

19. Filomeno R, Ikeda A, Maruyama K, Wada H, Tanigawa T. Excessive daytime sleepiness and alcohol consumption among commercial drivers. Occup Med Oxf Engl. 2019;69(6):406–11.

20. Koutsourelakis I, Perraki E, Bonakis A, Vagiakis E, Roussos C, Zakynthinos S. Determinants of subjective sleepiness in suspected obstructive sleep apnoea. J Sleep Res. 2008;17(4):437–43.

21. Bixler EO, Vgontzas AN, Lin HM, Calhoun SL, Vela-Bueno A, Kales A. Excessive daytime sleepiness in a general population sample: the role of sleep apnea, age, obesity, diabetes, and depression. J Clin Endocrinol Metab. 2005;90(8):4510–5.

22. Basta M, Lin HM, Pejovic S, Sarrigiannidis A, Bixler E, Vgontzas AN. Lack of regular exercise, depression, and degree of apnea are predictors of excessive daytime sleepiness in patients with sleep apnea: sex differences. J Clin Sleep Med. 2008;4(1):19–25.

23. Hwang H, Kim KM, Yun CH, Yang KI, Chu MK, Kim WJ. Sleep state of the elderly population in Korea: Nationwide cross-sectional population-based study. Front Neurol. 2022;13:1095404.

24. Fernandez-Mendoza J, Vgontzas AN, Kritikou I, Calhoun SL, Liao D, Bixler EO. Natural history of excessive daytime sleepiness: role of obesity, weight loss, depression, and sleep propensity. Sleep. 2015;38(3):351–60.

25. Ohayon MM, Carskadon MA, Guilleminault C, Vitiello MV. Meta-analysis of quantitative sleep parameters from childhood to old age in healthy individuals: developing normative sleep values across the human lifespan. Sleep. 2004;27(7):1255–73.

26. Stenuit P, Kerkhofs M. Age modulates the effects of sleep restriction in women. Sleep. 2005;28(10):1283–8.

